# Dietary Inflammatory Potential, Dietary Diversity and Micronutrient Adequacy Are Associated with Rheumatoid Arthritis and Autoantibody Burden: A Case-Control Study from India

**DOI:** 10.1101/2025.11.18.25340542

**Authors:** Hemant Mahajan, Sailaja Pothana, Phani Kumar, Janhavi Pawar, Srinivas K Ranga, Aditi Deshpande, B R Nikhita, Phani Kumar, Venkat Raji Reddy, Srikant Perugu, JJ Babu Geddam, Liza Rajasekhar, Devraj Parasannanavar

## Abstract

**Background:** Rheumatoid arthritis (RA) is a chronic autoimmune disease influenced by genetic and environmental factors, including diet. We examined the associations of dietary inflammatory potential, dietary diversity, and micronutrient adequacy with RA and RA-related autoantibodies in an urban Indian population.

**Methods:** In this unmatched case-control study, 301 newly diagnosed RA cases and 314 healthy controls from Hyderabad, India, were enrolled. Dietary intake was assessed using a validated 131-item food frequency questionnaire (FFQ). Dietary inflammatory potential was estimated using the Dietary Inflammatory Index (DII), while dietary quality was assessed using Dietary Diversity Score (DDS) and mean probability of micronutrient adequacy (MPA). Rheumatoid factor (RF) and anti-cyclic citrullinated peptide (anti-CCP) antibodies were measured by ELISA. Multivariable regression models adjusted for age, sex, and BMI were used to assess associations. Population attributable fractions (PAFs) were estimated under a theoretical causal framework.

**Results:** Compared with controls, RA cases had lower micronutrient adequacy (median MPA: 18.9% vs. 32.9%), lower dietary diversity (High DDS 50.2% vs. 74.2%), and higher dietary inflammatory scores (median DII: 2.18 vs. 0.37). Each 1-unit increase in DII was associated with higher odds of RA (AOR 1.17; 95% CI: 1.10, 1.24), whereas each 1-unit increase in DDS (AOR 0.73; 95% CI: 0.66, 0.81) and each 10% increase in MPA (AOR 0.88; 95% CI: 0.83, 0.94) were associated with lower odds of RA. High DDS and adequate micronutrient adequacy were also associated with lower RF and anti-CCP levels. Estimated PAFs for higher DII, low DDS, and nutrient inadequacy were 43.8%, 39.5%, and 37.5%, respectively.

**Conclusions:** Poorer dietary quality, lower micronutrient adequacy, and a more pro-inflammatory dietary profile were associated with higher odds of RA and greater autoantibody burden.

## INTRODUCTION

Rheumatoid arthritis (RA) is a chronic systemic autoimmune disease characterized by persistent synovial inflammation, progressive joint destruction, disability, and increased mortality. It affects approximately 0.5–1% of the global population, occurs two to three times more frequently in women than men, and commonly manifests during middle adulthood. [1,2,3]. The pathogenesis of RA involves genetic and environmental factors, with the HLA-DRB1 gene being strongly associated with disease susceptibility [4,5,6]. Environmental triggers, including cigarette smoking, infections, and dietary factors, interact with genetic susceptibility to influence disease onset and progression through immune dysregulation and inflammatory pathways[7,8,9,10]. Dietary intake plays a critical role in modulating inflammation, a central process in RA pathophysiology, through its effects on immune regulation, oxidative stress, gut microbiota, and inflammatory cytokine production [10–12].

Diets with anti-inflammatory properties, particularly the Mediterranean diet, have been associated with reduced disease activity, improved physical function, and symptom relief in patients with RA, whereas pro-inflammatory dietary patterns characterized by high intakes of processed foods, refined carbohydrates, saturated fats, and red meat (Western-style diets) may promote systemic inflammation and contribute to disease progression [10,11,13,14].Rapid urbanization and nutrition transition in India have resulted in increased consumption of energy-dense and animal-source foods, potentially contributing to chronic inflammatory conditions [15,16]. Western dietary patterns characterized by high intakes of saturated and trans fats, refined carbohydrates, processed foods, and an unfavorable omega-6:omega-3 fatty acid ratio promote systemic inflammation and have been associated with an increased risk of rheumatoid arthritis [10,11,17,18]. While dietary influences on inflammation in Western populations are well-documented, there is a paucity of research on the role of traditional Indian dietary patterns in RA-related inflammation. Macronutrients and micronutrients modulate RA pathophysiology by influencing immune function, oxidative stress, cytokine production, and other inflammatory pathways [10, 14,19].

Higher dietary fibre intake is inversely associated with inflammatory markers such as CRP, IL-6, and TNF-α, whereas Western dietary patterns characterized by low fibre and high consumption of processed and animal-derived foods are associated with greater systemic inflammation (10,20,21,22).Micronutrients, including antioxidant vitamins (C, E, and B-complex) and trace minerals such as magnesium, zinc, and selenium, modulate immune responses and oxidative stress, thereby exerting antioxidant and anti-inflammatory effects that may influence RA pathophysiology [10, 23,24,25,26,27]. Vitamin D and calcium are essential for maintaining bone health, while vitamin D also plays a key role in modulating innate and adaptive immune responses through regulation of inflammatory pathways[28,29,30].

Nutritional deficiencies, commonly observed in individuals with RA due to reduced dietary intake, altered nutrient metabolism, chronic inflammation, and medication-related effects, may aggravate systemic inflammation, increase oxidative stress, contribute to fatigue and functional impairment, and reduce health-related quality of life (10.29,30).Comprehensive tools like the Dietary Diversity Score (DDS) assess overall dietary quality and predict micronutrient adequacy [31,32], while the Dietary Inflammatory Index (DII) quantifies the inflammatory potential of a diet [12]. Existing research, however, primarily focuses on isolated dietary components or nutrients. Very few studies and none from India have explored the role of DII on RA [33,34,35]. The combined effects of dietary adequacy, diversity, and inflammatory potential on RA-related inflammation and progression remain underexplored, particularly in the context of Indian diets.

Despite the established role of diet in modulating inflammation, limited evidence exists on the dietary patterns, nutrient adequacy, and inflammatory potential of diets in RA patients in the Indian context. This study aims to evaluate nutritional adequacy, dietary diversity, and dietary inflammatory index among RA patients compared to healthy controls. By correlating these dietary factors with inflammatory markers, the study seeks to elucidate the relationship between diet quality, inflammatory potential, and RA pathogenesis. We hypothesized that higher DII and lower DDS and MPA would be associated with increased odds of RA, and that these dietary measures would be associated with higher RF and anti-CCP concentrations.

## METHODOLOGY

### Study setting and population

This unmatched case-control study was conducted in Hyderabad, India, and approved by the Institutional Ethics Committees of the National Institute of Nutrition (NIN IEC: 03/01/2014) and Nizam’s Institute of Medical Sciences (NIMS IEC: EC/NIMS/1665/2015; PBAC No. 1122/15). A total of 615 participants aged 18-55 years were enrolled, including 301 newly diagnosed RA cases and 314 healthy controls. Participants were recruited between 16 th June 2017 and 31 st December 2020, and written informed consent was obtained from all participants before enrolment. All participants were of Indian ethnicity and were recruited from Hyderabad and surrounding urban areas of Telangana State, India.

RA cases were diagnosed according to the 2010 ACR/EULAR classification criteria and confirmed by a Rheumatologist. Definite RA was defined by the presence of synovitis in at least one joint and a total ACR/EULAR score of ≥6/10. Controls were selected from schools, institutions, and private organizations in Hyderabad because recruitment of family/relative controls was not feasible. Frequency matching by age (±5 years) and sex was used at recruitment. Healthy controls were free from major chronic conditions, including type 2 diabetes, hypertension, PCOD, severe anaemia, thyroid dysfunction, and autoimmune diseases, and were not pregnant or lactating. For both cases and controls, exclusion criteria included juvenile RA, seronegative spondyloarthritis, inflammatory bowel disease, celiac disease, malabsorption disorders, and current use of antibiotics, probiotics, or immunosuppressive treatment. Participants with BMI <18.5 kg/m² or >30 kg/m² were also excluded.

### Anthropometric measurements and Dietary Assessment

The body weight was determined to the nearest 0.1 kg, they were measured using a SECA scale, and standing height was measured using a stadiometer with a precision of 0.5 cm. BMI was calculated for each individual, a value derived from a subject’s weight and height. BMI = weight (kg) **÷** height (m^2^). The dietary intake of subjects was obtained by using a semi-quantitative food frequency questionnaire (FFQ) including 131 dietary items divided into 13 food groups based on food classification according to the Indian Food Composition Tables (IFCT-2017) (36). To minimize errors while assessing subjects’ dietary intake, the interview process started by asking the subjects to recall all the food items consumed before the onset of the disease, listing as every day, weekly, once a month, and yearly. Based on the 12 standardized cups (ranging from 30 ml to 1400 ml) each participant’s intake was converted into weight equivalents (mg or g). The amount of daily food intake was calculated from the FFQ according to the following formula: Quantity of raw food ingredients consumed = (Quantity of raw food ingredient cooked **÷** Total Quantity of cooked food) **×** Quantity of cooked food consumed.

### Estimation of Nutrient Adequacy

Macronutrient adequacy was assessed using the Acceptable Macronutrient Distribution Range (AMDR) approach. For each participant, the percentage of total daily energy contributed by carbohydrate, fat, and protein was calculated and compared with the respective AMDR ranges.

Micronutrient adequacy was assessed using the probability of adequacy (PA) approach based on the Estimated Average Requirement (EAR), which is recommended for population-level assessment of dietary adequacy.

PA was calculated for 10 micronutrients: iron, zinc, thiamine, riboflavin, niacin, vitamin B12, vitamin B6, vitamin C, vitamin A, and folate, for which the required EAR values and corresponding coefficient of variation (CV)-based requirement standard deviation (SD) inputs were available and applied in our analysis. Micronutrients without the required inputs for the PA approach were not included in the Mean Probability of Adequacy (MPA) calculation.

For each micronutrient, the requirement SD was derived using the formula: SD = CV × EAR. The PA for each nutrient was then estimated using the EAR values provided by ICMR-NIN and the standard normal distribution function (NORM.S.DIST) in Microsoft Excel, representing the probability that an individual’s intake met the nutrient requirement. The Mean Probability of Adequacy (MPA) was calculated as the arithmetic mean of the probability of adequacy (PA) values for the 10 selected micronutrients. An MPA <0.5 was used as an operational criterion to indicate overall micronutrient inadequacy in the study population [32,37,38,39].

### Assessment of Dietary Diversity

Dietary Diversity Score (DDS) is a measure used to evaluate how diverse and nutritious an individual’s diet is. It shows the range of different food groups consumed within a specific time frame. Higher DDS values indicate better nutrient intake and overall health outcomes. Based on recent dietary guidelines 2024, the diet is divided into 10 food groups: cereals and millets, pulses, vegetables, nuts and oil seeds, green leafy vegetables, fruits, dairy, roots and tubers, flesh food and spices, and condiments. A score of 1 was assigned if a food group was consumed and 0 otherwise. The overall Dietary Diversity Score (DDS) was calculated by summing the scores across all food groups. Participants consuming fewer than five food groups (DDS <5) were classified as having low dietary diversity, whereas those consuming five or more food groups (DDS ≥5) were classified as having high dietary diversity [31].

### Assessment of Dietary Inflammatory Index (DII) Score

The FFQ-derived dietary data were utilized to compute the DII scores for each participant. A detailed explanation of the development and construct validation of the DII has been documented elsewhere. Following the approach described by Shivappa et al. [12], a counting algorithm was applied, based on an extensive literature review assessing the impact of diet on inflammatory biomarkers, including IL-4, TNF-α, IL-1β, IL-6, IL-10, and CRP. In this methodology, a total of 45 food parameters, encompassing both micronutrients and macronutrients, were assigned a score of (-1) for anti-inflammatory effects, (+1) for pro-inflammatory effects, or (0) if they had no impact on these inflammatory biomarkers.

In this study, dietary data from 36 food parameters collected through the FFQ were available for DII calculation. These parameters included: *Energy, Iron, Protein, Zinc, Carbohydrates, Thiamine, Fats, Riboflavin, MUFA, Vitamin B12, PUFA, Vitamin C, Trans fats, Vitamin A, Fiber, Vitamin D3, Folic acid, Vitamin B6, β-carotene, Total cholesterol, Vit E, Magnesium, Niacin, Selenium, Garlic, n-3 Fatty acids, Ginger, n-6 Fatty acids, Turmeric, Onion, Flavonones, Saturated Fat, Isoflavones, Pepper, Flavones, Flavan-3-ol* (Supplementary table 1). Each food constituent was assigned a DII score based on a global standard database. This score was calculated by subtracting the global mean from the reported intake and dividing by the standard deviation, resulting in a Z-score. To account for the right-skewed distribution commonly observed in dietary data, the Z-score was converted into a centered percentile score. Each food parameter’s centered percentile score was then multiplied by its respective food parameter-specific inflammatory effect score, as determined from literature reviews.

The final DII score for each participant was obtained by summing all food parameter-specific DII scores, providing an overall measure of dietary inflammatory potential.

### Antibody Measurement

Non-fasting venous blood samples (approximately 10 mL per participant) were collected by trained phlebotomists using aseptic techniques and sterile vacutainer tubes with appropriate anticoagulants (EDTA for plasma; serum separator tubes for serum). Serum and plasma were separated within 2 hours of collection by centrifugation (1,500×g for 10 minutes) and stored in aliquots at −80°C until laboratory analysis (within 6 months). Serum Rheumatoid Factor (RF) and Plasma anti-cyclic citrullinated peptide (Anti-CCP) levels were performed using ELISA technique as per manufacturer instructions (RF was detected by IgM RF ELISA, APSLABS India), values were interpreted in IU/ml. Anti-CCP antibodies were detected by ELISA using commercially available kits (DRG, Germany) and values were interpreted in U/mL.

### Statistical Analysis

Continuous data were expressed as median and interquartile range (IQR) and compared between cases and controls using the Mann-Whitney U test. Categorical data were presented as frequency and percentage and analyzed using Pearson’s Chi-square test to assess differences between the two groups. To evaluate the independent associations of DDS, DII, and MPA with RA, we conducted separate unconditional logistic regression models for each dietary variable, adjusting for age, sex, and BMI. Unconditional logistic regression was chosen because complete frequency matching based on age and sex was not fully achieved. To further minimize the risk of residual confounding, we included age, sex, and BMI as covariates in all models, ensuring appropriate adjustment for these confounders. Multiple linear regression models adjusting for age, sex and BMI were used to assess the independent associations of DDS, DII, and MPA with Anti-CCP and RF antibodies. Additionally, we created the tertiles of DII and assessed its relationship with RA, Anti-CCP and RF antibodies using appropriate regression models. The population attributable fraction (PAF) was calculated for DII, DDS (DDS<5.0 vs. DDS ≥5.0), and MPA (MPA <0.5 vs. MPA ≥0.5). For PAF estimation related to DII, tertiles 2 and 3 were combined and compared to tertile 1. To calculate PAF for odds ratio (OR) >1.0 and OR <1.0, we used Levin’s formula: [PAF= P**_exposed_**\*(OR - 1) **÷** (1 + P**_exposed_** *(1-OR))] and [PAF= P**_exposed_**\*(1 - OR) **÷** (1-P**_exposed_** *(1-OR))], respectively. We used Wald Method to calculate 95% CI of PAF. All tests were two-sided, and p-values <0.05 was considered statistically significant to account for multiple testing.

## RESULTS

### Participant’s Characteristics

The median age of the participants was 38.0 years (IQR: 32.0–45.0), with cases being significantly older than controls (41.0 vs. 35.0 years, p < 0.001). Females constituted the majority of the study population (88.6%), with comparable proportions in cases (87.0%) and controls (90.1%) (p = 0.486). The median BMI was significantly lower among cases compared to controls (23.14 vs. 25.62 kg/m², p < 0.001). Cases reported consistently lower intake of macronutrients compared to controls, including energy (1908.22 vs. 2138.95 kcal), protein (42.75 vs. 50.93 g), carbohydrates (235.35 vs. 266.20 g), fats (71.64 vs. 84.54 g), and fibre (21.38 vs. 28.00 g), all p < 0.001. Similarly, the median intake of several micronutrients was significantly lower among cases, including iron (8.85 vs. 12.25 mg), zinc (5.91 vs. 6.53 mg), thiamine (0.49 vs. 0.68 mg), riboflavin (0.37 vs. 0.50 mg), niacin (7.48 vs. 8.84 mg), vitamin B6 (0.30 vs. 0.43 mg), vitamin C (72.52 vs. 98.22 mg), vitamin A (390.94 vs. 509.40 RE), beta-carotene (350.89 vs. 577.53 mcg), folate (180.89 vs. 238.68 mcg), selenium (2.35 vs. 4.52 mcg), vitamin E (0.65 vs. 1.00 mg), and magnesium (65.30 vs. 101.89 mg), all p < 0.001. However, no significant differences were observed for vitamin B12 and vitamin D intake.

Additionally, cases had lower intake of total cholesterol (16.88 vs. 30.11 mg) and trans fats (0.89 vs. 1.60 g) compared to controls (p < 0.001). Intake of fatty acids, including MUFA (0.12 vs. 0.26 g), PUFA (0.35 vs. 0.50 g), n-3 fatty acids (0.10 vs. 0.13 g), and n-6 fatty acids (0.24 vs. 0.40 g), was also significantly lower among cases. Furthermore, consumption of bioactive compounds such as flavones, flavanones, flavan-3-ols, and isoflavones, as well as spices like onion, ginger, turmeric, and pepper, was significantly lower in cases compared to controls (Table 1).

**Table 1.**
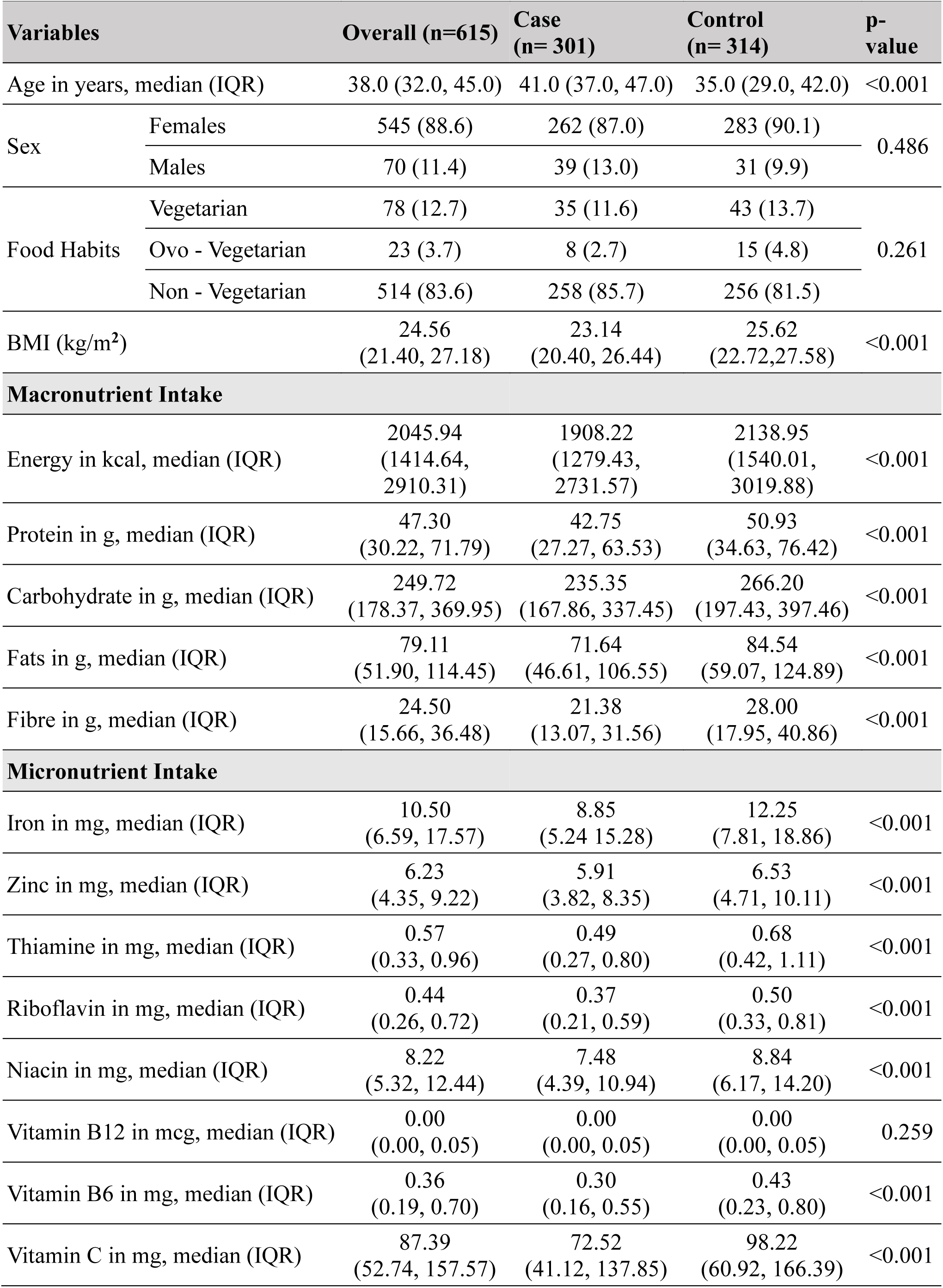
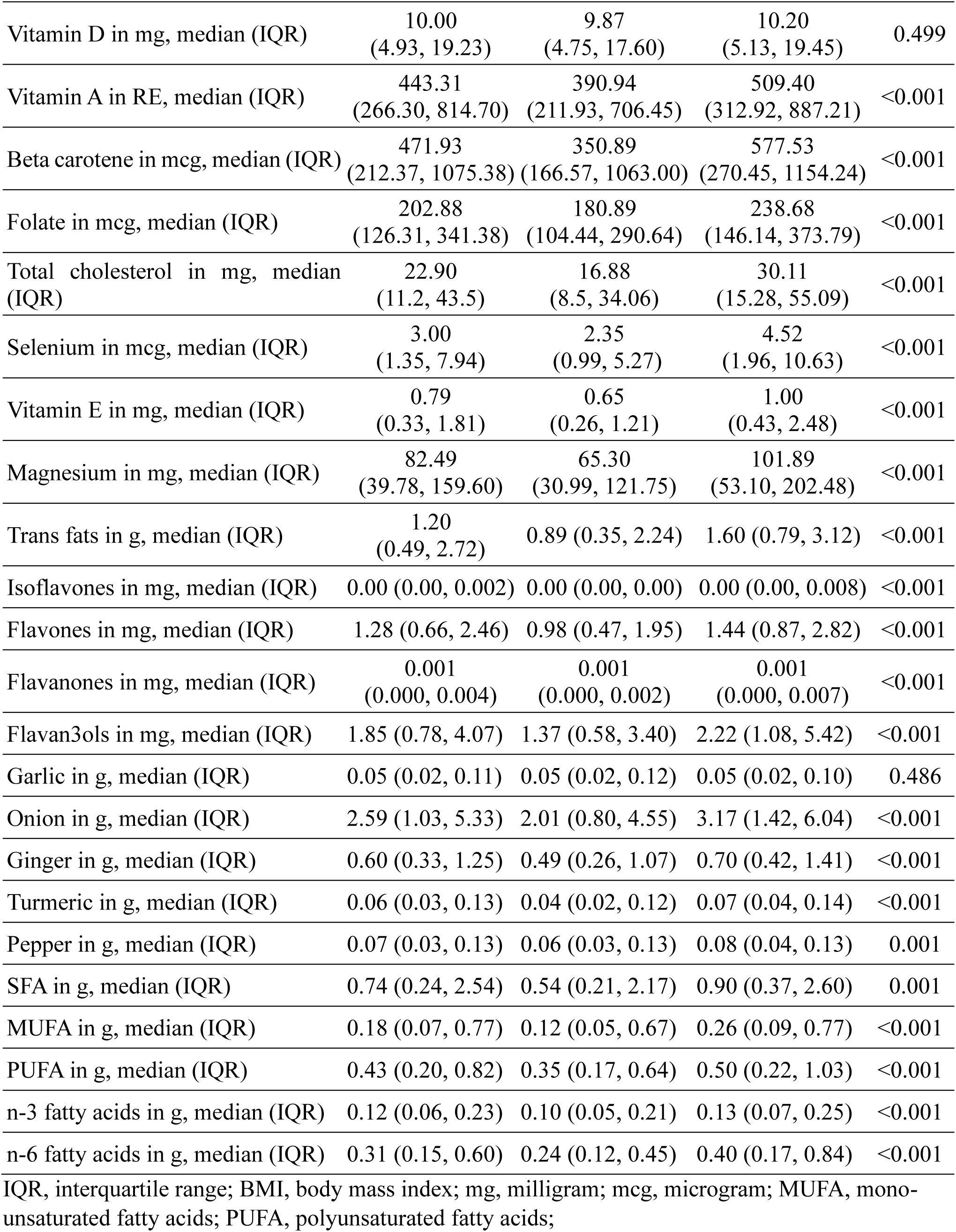
Demographic, body mass index and nutrient consumption distribution across participants.

| Variables |  | Overall (n=615) | Case (n= 301) | Control (n= 314) | p-value |
| --- | --- | --- | --- | --- | --- |
| Age in years, median (IQR) |  | 38.0 (32.0, 45.0) | 41.0 (37.0, 47.0) | 35.0 (29.0, 42.0) | <0.001 |
| Sex | Females | 545 (88.6) | 262 (87.0) | 283 (90.1) | 0.486 |
|  | Males | 70 (11.4) | 39 (13.0) | 31 (9.9) |  |
| Food Habits | Vegetarian | 78 (12.7) | 35 (11.6) | 43 (13.7) | 0.261 |
|  | Ovo - Vegetarian | 23 (3.7) | 8 (2.7) | 15 (4.8) |  |
|  | Non - Vegetarian | 514 (83.6) | 258 (85.7) | 256 (81.5) |  |
| BMI (kg/m<sup>2</sup>) |  | 24.56<br>(21.40, 27.18) | 23.14<br>(20.40, 26.44) | 25.62<br>(22.72,27.58) | <0.001 |
| **Macronutrient Intake** |  |  |  |  |  |
| Energy in kcal, median (IQR) |  | 2045.94<br>(1414.64, 2910.31) | 1908.22<br>(1279.43, 2731.57) | 2138.95<br>(1540.01, 3019.88) | <0.001 |
| Protein in g, median (IQR) |  | 47.30<br>(30.22, 71.79) | 42.75<br>(27.27, 63.53) | 50.93<br>(34.63, 76.42) | <0.001 |
| Carbohydrate in g, median (IQR) |  | 249.72<br>(178.37, 369.95) | 235.35<br>(167.86, 337.45) | 266.20<br>(197.43, 397.46) | <0.001 |
| Fats in g, median (IQR) |  | 79.11<br>(51.90, 114.45) | 71.64<br>(46.61, 106.55) | 84.54<br>(59.07, 124.89) | <0.001 |
| Fibre in g, median (IQR) |  | 24.50<br>(15.66, 36.48) | 21.38<br>(13.07, 31.56) | 28.00<br>(17.95, 40.86) | <0.001 |
| **Micronutrient Intake** |  |  |  |  |  |
| Iron in mg, median (IQR) |  | 10.50<br>(6.59, 17.57) | 8.85<br>(5.24 15.28) | 12.25<br>(7.81, 18.86) | <0.001 |
| Zinc in mg, median (IQR) |  | 6.23<br>(4.35, 9.22) | 5.91<br>(3.82, 8.35) | 6.53<br>(4.71, 10.11) | <0.001 |
| Thiamine in mg, median (IQR) |  | 0.57<br>(0.33, 0.96) | 0.49<br>(0.27, 0.80) | 0.68<br>(0.42, 1.11) | <0.001 |
| Riboflavin in mg, median (IQR) |  | 0.44<br>(0.26, 0.72) | 0.37<br>(0.21, 0.59) | 0.50<br>(0.33, 0.81) | <0.001 |
| Niacin in mg, median (IQR) |  | 8.22<br>(5.32, 12.44) | 7.48<br>(4.39, 10.94) | 8.84<br>(6.17, 14.20) | <0.001 |
| Vitamin B12 in mcg, median (IQR) |  | 0.00<br>(0.00, 0.05) | 0.00<br>(0.00, 0.05) | 0.00<br>(0.00, 0.05) | 0.259 |
| Vitamin B6 in mg, median (IQR) |  | 0.36<br>(0.19, 0.70) | 0.30<br>(0.16, 0.55) | 0.43<br>(0.23, 0.80) | <0.001 |
| Vitamin C in mg, median (IQR) |  | 87.39<br>(52.74, 157.57) | 72.52<br>(41.12, 137.85) | 98.22<br>(60.92, 166.39) | <0.001 |

|  |  |  |  |  |
| --- | --- | --- | --- | --- |
| Vitamin D in mg, median (IQR) | 10.00<br>(4.93, 19.23) | 9.87<br>(4.75, 17.60) | 10.20<br>(5.13, 19.45) | 0.499 |
| Vitamin A in RE, median (IQR) | 443.31<br>(266.30, 814.70) | 390.94<br>(211.93, 706.45) | 509.40<br>(312.92, 887.21) | <0.001 |
| Beta carotene in mcg, median (IQR) | 471.93<br>(212.37, 1075.38) | 350.89<br>(166.57, 1063.00) | 577.53<br>(270.45, 1154.24) | <0.001 |
| Folate in mcg, median (IQR) | 202.88<br>(126.31, 341.38) | 180.89<br>(104.44, 290.64) | 238.68<br>(146.14, 373.79) | <0.001 |
| Total cholesterol in mg, median (IQR) | 22.90<br>(11.2, 43.5) | 16.88<br>(8.5, 34.06) | 30.11<br>(15.28, 55.09) | <0.001 |
| Selenium in mcg, median (IQR) | 3.00<br>(1.35, 7.94) | 2.35<br>(0.99, 5.27) | 4.52<br>(1.96, 10.63) | <0.001 |
| Vitamin E in mg, median (IQR) | 0.79<br>(0.33, 1.81) | 0.65<br>(0.26, 1.21) | 1.00<br>(0.43, 2.48) | <0.001 |
| Magnesium in mg, median (IQR) | 82.49<br>(39.78, 159.60) | 65.30<br>(30.99, 121.75) | 101.89<br>(53.10, 202.48) | <0.001 |
| Trans fats in g, median (IQR) | 1.20<br>(0.49, 2.72) | 0.89 (0.35, 2.24) | 1.60 (0.79, 3.12) | <0.001 |
| Isoflavones in mg, median (IQR) | 0.00 (0.00, 0.002) | 0.00 (0.00, 0.00) | 0.00 (0.00, 0.008) | <0.001 |
| Flavones in mg, median (IQR) | 1.28 (0.66, 2.46) | 0.98 (0.47, 1.95) | 1.44 (0.87, 2.82) | <0.001 |
| Flavanones in mg, median (IQR) | 0.001<br>(0.000, 0.004) | 0.001<br>(0.000, 0.002) | 0.001<br>(0.000, 0.007) | <0.001 |
| Flavan3ols in mg, median (IQR) | 1.85 (0.78, 4.07) | 1.37 (0.58, 3.40) | 2.22 (1.08, 5.42) | <0.001 |
| Garlic in g, median (IQR) | 0.05 (0.02, 0.11) | 0.05 (0.02, 0.12) | 0.05 (0.02, 0.10) | 0.486 |
| Onion in g, median (IQR) | 2.59 (1.03, 5.33) | 2.01 (0.80, 4.55) | 3.17 (1.42, 6.04) | <0.001 |
| Ginger in g, median (IQR) | 0.60 (0.33, 1.25) | 0.49 (0.26, 1.07) | 0.70 (0.42, 1.41) | <0.001 |
| Turmeric in g, median (IQR) | 0.06 (0.03, 0.13) | 0.04 (0.02, 0.12) | 0.07 (0.04, 0.14) | <0.001 |
| Pepper in g, median (IQR) | 0.07 (0.03, 0.13) | 0.06 (0.03, 0.13) | 0.08 (0.04, 0.13) | 0.001 |
| SFA in g, median (IQR) | 0.74 (0.24, 2.54) | 0.54 (0.21, 2.17) | 0.90 (0.37, 2.60) | 0.001 |
| MUFA in g, median (IQR) | 0.18 (0.07, 0.77) | 0.12 (0.05, 0.67) | 0.26 (0.09, 0.77) | <0.001 |
| PUFA in g, median (IQR) | 0.43 (0.20, 0.82) | 0.35 (0.17, 0.64) | 0.50 (0.22, 1.03) | <0.001 |
| n-3 fatty acids in g, median (IQR) | 0.12 (0.06, 0.23) | 0.10 (0.05, 0.21) | 0.13 (0.07, 0.25) | <0.001 |
| n-6 fatty acids in g, median (IQR) | 0.31 (0.15, 0.60) | 0.24 (0.12, 0.45) | 0.40 (0.17, 0.84) | <0.001 |

IQR, interquartile range; BMI, body mass index; mg, milligram; mcg, microgram; MUFA, mono-unsaturated fatty acids; PUFA, polyunsaturated fatty acids;

### Nutrient adequacy

According to AMDR guidelines, macronutrient adequacy was suboptimal across both groups, with consistently lower adequacy observed among cases. A significantly lower proportion of cases met the carbohydrate AMDR compared to controls (41.5% vs. 50.3%, p = 0.029). Protein adequacy was also low in both groups, with only 19.9% of cases meeting the AMDR compared to 28.0% of controls (p = 0.019). Similarly, fat intake within the AMDR was slightly but significantly lower among cases (80.0%) compared to controls (86.3%) (p = 0.049). Energy adequacy was also lower in cases (49.5%) relative to controls (57.6%) (p = 0.043).Fibre intake was notably lower among cases, with median intake at 76.34% of EAR (IQR: 46.68–115.84) compared to 100.37% (IQR: 62.17–147.57) in controls (p < 0.001). Correspondingly, a significantly smaller proportion of cases met fibre adequacy (71.1%) compared to controls (88.5%) (p < 0.001).

The PA for several micronutrients was generally low, particularly among cases. Median PA for iron was substantially lower in cases compared to controls (0.13 vs. 0.99, p < 0.001). Similarly, zinc (0.00 vs. 0.01, p < 0.001) and niacin (0.01 vs. 0.04, p < 0.001) showed significantly lower adequacy among cases. For thiamine, riboflavin, vitamin B6, and vitamin B12, the median PA was near zero in both groups, indicating widespread inadequacy.In contrast, the probability of adequacy for vitamin C was high in both groups but remained significantly lower among cases (0.99 vs. 1.00, p < 0.001). Vitamin A adequacy was also significantly lower in cases compared to controls (0.05 vs. 0.28, p < 0.001), while folate adequacy was uniformly high across both groups.Overall, the median MPA was significantly lower among cases compared to controls (18.88% vs. 32.87%, p < 0.001). Correspondingly, only 21.3% of cases were nutritionally adequate compared to 34.7% of controls (p < 0.001), highlighting poorer overall nutrient adequacy among cases (Table 2). Although several micronutrients showed median PA values close to zero, meaningful between-group differences remained evident for MPA and selected micronutrients.

**Table 2.** Acceptable Macronutrient Distribution Range and probability of Adequacy of micronutrients.

|  | **Overall (n=615)** | **Case (n= 301)** | **Control (n= 314)** | **p-value** |
| --- | --- | --- | --- | --- |
| **Acceptable Macronutrient Distribution Range** |  |  |  |  |
| Protein AMDR (Adequate), n(%) | 148 (24.1) | 60 (19.9) | 88 (28.0) | 0.019 |
| Fat AMDR (Adequate), n(%) | 513 (83.4) | 242 (80.0) | 271 (86.3) | 0.049 |
| Carbohydrate AMDR (Adequate), n(%) | 283 (46.0) | 125 (41.5) | 158 (50.3) | 0.029 |
| Energy status (Adequate), n(%) | 330 (53.7) | 149 (49.5) | 181 (57.6) | 0.043 |
| Fiber consumption (in % EAR), median (IQR) | 85.37<br>(56.14, 128.55) | 76.34<br>(46.68, 115.84) | 100.37<br>(62.17, 147.57) | <0.001 |
| Fiber consumption status (Adequate), n(%) | 492 (80.0) | 214 (71.1) | 278 (88.5) | <0.001 |
| **Probability of Adequacy of Micronutrients** |  |  |  |  |
| Iron, median (IQR) | 0.69<br>(0.00, 1.00) | 0.13<br>(0.00, 1.00) | 0.99<br>(0.01, 1.00) | <0.001 |
| Zinc, median (IQR) | 0.01<br>(0.00, 0.76) | 0.00<br>(0.00, 0.38) | 0.01<br>(0.00, 0.96) | <0.001 |
| Thiamine, median (IQR) | 0.00<br>(0.00, 0.33) | 0.00<br>(0.00, 0.04) | 0.00<br>(0.00, 0.88) | - |
| Riboflavin, median (IQR) | 0.00<br>(0.00, 0.00) | 0.00<br>(0.00, 0.00) | 0.00<br>(0.00, 0.01) | - |
| Niacin, median (IQR) | 0.02<br>(0.00, 0.60) | 0.01<br>(0.00, 0.28) | 0.04<br>(0.00, 0.89) | <0.001 |
| Vitamin B12, median (IQR) | 0.00<br>(0.00, 0.00) | 0.00<br>(0.00, 0.00) | 0.00<br>(0.00, 0.00) | - |
| Vitamin B6, median (IQR) | 0.00<br>(0.00, 0.00) | 0.00<br>(0.00, 0.00) | 0.00<br>(0.00, 0.00) | - |
| Vitamin C, median (IQR) | 1.00<br>(0.11, 1.00) | 0.99<br>(0.00, 1.00) | 1.00<br>(0.56, 1.00) | <0.001 |
| Vitamin A, median (IQR) | 0.12<br>(0.01, 0.98) | 0.05<br>(0.00, 0.86) | 0.28<br>(0.01, 1.00) | <0.001 |
| Folate, median (IQR) | 1.00<br>(1.00, 1.00) | 1.00<br>(1.00, 1.00) | 1.00<br>(1.00, 1.00) | - |
| MPA in %, median (IQR) | 24.24<br>(4.63, 55.89) | 18.88<br>(0.72, 43.18) | 32.87<br>(12.51, 63.01) | <0.001 |
| MPA status (Adequate), n(%) | 173 (28.1) | 64 (21.3) | 109 (34.7) | <0.001 |

AMDR, Acceptable Macronutrient Distribution Range; IQR, interquartile range; MPA, mean probability of adequacy; n, frequency; %, percentage; MPA, mean probability of adequacy; EAR, Estimated Average Requirement;

**Table 3.** Distribution of mean probability of adequacy of micronutrients, dietary score and dietary inflammation across participants.

|  | **Overall (n=615)** | **Case (n= 301)** | **Control (n= 314)** | **p-value** |
| --- | --- | --- | --- | --- |
| DDS, median (IQR) | 5.0<br>(4.0, 6.0) | 5.0<br>(3.0, 6.0) | 5.0<br>(4.0, 7.0) | <0.001 |
| DDS status (High), n (%) | 384 (62.4) | 151 (50.2) | 233 (74.2) | <0.001 |
| DII, median (IQR) | 1.52<br>(-1.57, 3.28) | 2.18<br>(-0.43, 3.81) | 0.37<br>(-2.19, 2.73) | <0.001 |
| Anti-CCP in EU/mL, median (IQR) | 15.8<br>(7.0, 100) | 98.6<br>(15.1, 647.9) | 9.2<br>(3.72, 16.3) | <0.001 |
| RF in U/mL, median (IQR) | 31.0<br>(12.36, 377.0) | 385.5<br>(139.0, 825.6) | 12.6<br>(7.3, 17.3) | <0.001 |

DDS, dietary diversity score; DDS score $\geq 5$ is considered as *High*; IQR, interquartile range; n, frequency; %, percentage; RF, Rheumatoid Factor; Anti-CCP, anti-cyclic citrullinated peptide;

**Table 4.** Association of dietary inflammation, dietary score with rheumatoid arthritis and associated antibodies.

| Variables | Rheumatoid Arthritis |  | Anti-CCP antibodies |  | RF antibodies |  |
| --- | --- | --- | --- | --- | --- | --- |
| | AOR (95% CI) | p-value | β-coeff (95% CI) | p-value | β-coeff (95% CI) | p-value |
| DII (per unit change) | 1.17<br>(1.10, 1.24) | <0.001 | 5.18<br>(-5.21, 15.58) | 0.328 | 7.91<br>(-4.20, 20.01) | 0.200 |
| DII Tertiles* |  |  | <0.001 |  | 0.497 | 0.585 |
| Tertile 1 | 1.00 | - | 0.00 | - | 0.00 | - |
| Tertile 2 | 1.86<br>(1.21, 2.85) | 0.005 | 76.01<br>(2.26, 149.76) | 0.043 | 52.71<br>(-33.44, 138.86) | 0.230 |
| Tertile 3 | 2.55<br>(1.65, 3.93) | <0.001 | 25.58<br>(-48.25, 99.41) | 0.497 | 23.99<br>(-62.25, 110.24) | 0.585 |
| DDS (per unit change) | 0.73<br>(0.66, 0.81) | <0.001 | -22.25<br>(-38.82, -5.69) | 0.009 | -11.25<br>(-30.65, 8.15) | 0.255 |
| DDS status<br>(High vs Low (ref)) | 0.37<br>(0.26, 0.54) | <0.001 | -36.99<br>(-99.39, 25.41) | 0.245 | -51.49<br>(-124.18, 21.20) | 0.165 |
| MPA in % (per 10% change) | 0.88<br>(0.83, 0.94) | <0.001 | -2.53<br>(-12.87, 7.80) | 0.630 | -6.15<br>(-18.18, 5.88) | 0.316 |
| MPA status<br>($\geq 0.5$ vs $< 0.5$ (ref)) | 0.54 (0.37, 0.80) | <0.001 | -20.48<br>(-87.91, 46.96) | 0.551 | -44.65<br>(-123.19, 33.88) | 0.265 |

AOR, adjusted odds ratio; CI, confidence interval; DII, dietary inflammatory index; DDS, dietary diversity score; IQR, interquartile range; n, frequency; %, percentage; RF, Rheumatoid Factor; Anti-CCP, anti-cyclic citrullinated peptide; MPA, mean probability of adequacy; ref, reference category; DDS score $\geq 5$ is considered as *High*; MPA $\geq 0.5$ considered as nutrient adequacy; Regression models were adjusted for age, sex and body mass index; \*p-trend across the tertiles of DII for all outcome variables.

### Dietary diversity and Dietary inflammation

The median DDS differed significantly between the cases and controls, with lower diversity observed among cases (5.0 [3.0, 6.0]) compared to controls (5.0 [4.0, 7.0]) (p <0.001). A smaller proportion of cases achieved a high DDS (≥5), with 50.2% classified as having high DDS compared to 74.2% among controls (p <0.001).

The DII scores ranged from -7.18 (maximally anti-inflammatory) to +4.93 (maximally pro-inflammatory). The median DII score was significantly higher among cases (2.18 [-0.43, 3.81]) compared to controls (0.37 [-2.19, 2.73]) (p <0.001).

Additionally, biomarkers of disease activity were markedly elevated among cases. Median anti-CCP levels were substantially higher in the cases (98.6 [15.1, 647.9] EU/mL) compared to controls (9.2 [3.72, 16.3] EU/mL). Similarly, RF levels were significantly elevated among cases (385.5 [139.0, 825.6] U/mL) relative to controls (12.6 [7.3, 17.3] U/mL) (p <0.001 for both comparisons).

After adjusting for age, sex, and BMI, each one-unit increase in the DII was associated with a-17% higher odds of RA [AOR (95% CI): 1.17 (1.10, 1.24)]. The odds of RA increased across tertiles of DII (p-trend <0.001), with participants in tertile 2 and tertile 3 having 1.86-fold [AOR (95% CI): 1.21, 2.85] and 2.55-fold [AOR (95% CI): 1.65, 3.93] higher odds of RA, respectively, compared to tertile 1. A one-unit increase in the DDS was associated with a 27% lower likelihood of RA [AOR (95% CI): 0.73 (0.66, 0.81)]. Participants with high DDS (≥5) had significantly lower odds of RA compared to those with low DDS, corresponding to a 63% reduction in odds [AOR (95% CI): 0.37 (0.26, 0.54)]. Each 10% increase in MPA of nutrients was associated with a 12% reduction in the odds of RA [AOR (95% CI): 0.88 (0.83, 0.94)]. Participants with adequate MPA (≥0.5) had 46% lower odds of RA compared to those with inadequate MPA [AOR (95% CI): 0.54 (0.37, 0.80)].

The PAFs indicated that 43.8% (24.6, 58.0) of RA cases could be attributed to higher DII (tertiles 2 and 3 vs. tertile 1), 39.5% (25.3, 49.9) to low DDS (<5 vs. ≥5), and 37.7% (14.7, 54.6) to inadequate MPA (<0.5 vs. ≥0.5), assuming causality.

Regarding antibody levels, DII (per unit increase) was not significantly associated with anti-CCP [β-coeff. (95% CI): 5.18 (-5.21, 15.58)] or RF levels [7.91 (-4.20, 20.01)]. However, compared to the lowest tertile, participants in DII tertile 2 had significantly higher anti-CCP levels [β-coeff. (95% CI): 76.01 (2.26, 149.76)], while no significant associations were observed for tertile 3 or for RF levels. Each one-unit increase in DDS was associated with significantly lower anti-CCP levels [β-coeff. (95% CI): -22.25 (-38.82, -5.69)], although its association with RF was not statistically significant. Similarly, high DDS status was not significantly associated with anti-CCP or RF levels. MPA (per 10% increase) and MPA status were not significantly associated with anti-CCP or RF antibody levels, although the direction of association suggested lower antibody levels with higher nutrient adequacy.

## DISCUSSION

Unlike previous studies that primarily focused on individual nutrients, dietary patterns, or DII alone, the present study simultaneously assessed dietary inflammatory potential, dietary diversity, micronutrient adequacy, and RA-specific autoantibodies. This integrated approach provides a broader evaluation of dietary quality and its potential relationship with autoimmune activity in RA.

This community-based unmatched case-control study examined the relationship between the dietary inflammation, diet diversity, micronutrient adequacy, and RA risk. The results indicate that RA cases had significantly higher DII scores, reflecting more pro-inflammatory diets, and lower DDS, indicating poor dietary diversity, compared to controls. Poor dietary quality may represent modifiable factors associated with RA risk and higher anti-CCP levels, whereas micronutrient inadequacy was associated with increased RA risk but showed no statistically significant association with RF or anti-CCP levels. Specifically, each one-unit increase in DII was associated with a 17% increase in RA odds, while a one-unit increase in DDS was linked to a 27% reduction in RA odds. Each 10% increase in MPA of nutrients was associated with a 12% reduction in the odds of RA. The PAF analyses indicate that 43.8% (24.6-58.0) of cases could be attributed to a higher DII, 39.5% (25.3-49.9) to lower DDS, and 37.7% (14.7-54.6) to inadequate nutrient intake (probability of nutrient adequacy <50%), assuming a causal relationship.

These findings highlight the significant role of diet in the development and progression of RA. Our findings align with previous studies that underscore the role of diet in modulating inflammation and RA risk. Gao et al. [33] reported higher odds of RA in individuals consuming pro-inflammatory diets, with the highest DII tertile associated with elevated disease risk. Similarly, Nayebi [34] and Tandorost [40] observed that higher DII scores correlated with increased serum inflammatory markers (e.g., hs-CRP, TNF-α) and worse clinical outcomes (e.g., DAS-28 score). Evidence from Japan [41] and the UK Biobank [42] further supports the association between higher DII scores and increased RA activity and inflammation. In Japan, the TOMORROW study reported that RA patients had significantly higher energy-adjusted DII (E-DII) scores compared to controls, suggesting a more pro-inflammatory dietary pattern. While the E-DII score itself was not directly linked to significant changes in disease activity, a shift towards a more anti-inflammatory diet was associated with the maintenance of low disease activity over a six-year follow-up period. These findings highlight the potential role of dietary inflammation in modulating RA progression [41].

Similarly, data from the UK Biobank revealed a significant association between diet, inflammation (measured by C-reactive protein levels), and RA, suggesting that higher DII scores are linked to increased RA activity and inflammation [42]. Conversely, anti-inflammatory dietary patterns characterized by high consumption of fruits, vegetables, whole grains, legumes, and omega-3 fatty acids have been shown to reduce systemic inflammation, improve disease activity, and alleviate clinical symptoms in patients with rheumatoid arthritis [10,11,13,14,43].

Moreover, studies on dietary patterns indicate that adherence to anti-inflammatory diets, such as Mediterranean or vegan diets, can reduce systemic inflammation and RA disease activity [44,45]. For example, randomized controlled trials have demonstrated that omega-3 supplementation reduces RA-related inflammation, while diets high in sugar, red meat, and saturated fats exacerbate disease progression [44,45,46]. Our study expands on these findings by showing the specific impact of dietary inadequacy and low DDS on RA risk and antibody levels.

A plausible alternative explanation for the observed associations could be reverse causation, whereby early RA symptoms (e.g., pain, stiffness, fatigue, reduced appetite, and difficulty preparing or consuming certain foods) may have influenced food choices and overall intake before or around diagnosis, rather than diet alone contributing to disease risk. RA-related symptom burden may lead to lower energy intake, reduced dietary diversity, and poorer micronutrient adequacy, which could partly explain the more pro-inflammatory dietary profile observed in cases. In addition, health-seeking behaviour after symptom onset may have prompted selective dietary changes or differential recall of “pre-disease” diet among cases compared with controls. Although we attempted to reduce this concern by recruiting newly diagnosed RA cases and assessing habitual intake using a validated interviewer-administered FFQ, these alternative explanations cannot be excluded in a case-control design and should be considered when interpreting the findings.

### Mechanisms Underlying DII and Rheumatoid Arthritis

The relationship between DII and RA can be attributed to multiple biological mechanisms. Pro-inflammatory dietary patterns rich in saturated fats, refined carbohydrates, and processed foods promote systemic inflammation by enhancing the production of pro-inflammatory cytokines, including interleukin-6 (IL-6) and tumor necrosis factor-α (TNF-α), which play central roles in RA pathogenesis [10,20]. These cytokines are central mediators of RA pathogenesis, promoting persistent synovial inflammation, pannus formation, cartilage degradation, bone erosion, and the maintenance of autoimmune responses [47,48].

High DII scores, reflecting poor dietary quality, can also exacerbate oxidative stress and impair the body’s antioxidant defence systems, further promoting inflammatory cascades.

Dietary patterns may influence RA pathogenesis through modulation of the gut microbiota. Pro-inflammatory dietary patterns are associated with gut dysbiosis, impaired intestinal barrier function, and heightened immune activation, whereas diets rich in dietary fiber, omega-3 fatty acids, and polyphenols promote microbial diversity, increase the production of anti-inflammatory metabolites such as short-chain fatty acids, and may contribute to reduced systemic inflammation and autoimmunity (10,11,17,18,43,49,50)

Micronutrient inadequacy, as observed in this study, may contribute to RA progression by increased oxidative stress and antioxidant defence mechanisms. Deficiencies in vitamins B6, C, and E, along with trace minerals such as zinc and selenium, have been associated with increased oxidative stress and heightened inflammatory responses in RA (10,19,23,24,25). Adequate intake of these micronutrients is critical for maintaining immune homeostasis and mitigating RA-associated inflammation.

### Public Health Significance

The findings of this study have important public health implications. RA is a chronic autoimmune disease with significant physical, emotional, and economic burdens. While pharmacological treatments are effective, they are often associated with adverse effects and high costs. Dietary interventions, on the other hand, offer a cost-effective, sustainable, and side-effect-free approach to reducing RA risk and managing disease activity. Promoting diets with low DII scores, rich in fruits, vegetables, whole grains, and omega-3 fatty acids can serve as a preventive strategy against RA and other inflammatory diseases. Public health campaigns should emphasize the importance of dietary diversity and adequate micronutrient intake in reducing inflammation and supporting immune health. Additionally, integrating dietary counselling into RA management programs could improve patient outcomes by complementing pharmacological treatments with lifestyle modifications. This study highlights the critical role of diet in RA onset and progression. A pro-inflammatory diet and poor dietary quality are significant risk factors for RA, while anti-inflammatory diets and adequate nutrient intake may help mitigate disease risk and severity. Future research should focus on longitudinal studies and clinical trials to further validate these findings and develop targeted dietary interventions for RA prevention and management.

### Strengths and Limitations

Our study has several strengths. First, the relatively large sample size (615 participants) allowed us to examine the association between dietary exposures and RA with reasonable precision while adjusting for key confounders. Second, diet was assessed using a validated FFQ adapted with locally collected recipes, which improves the relevance of dietary intake estimates in this setting. The simultaneous evaluation of DII, DDS, and MPA has rarely been reported in South Asian populations.

In addition, RA cases were clinically diagnosed, and both anti-CCP and RF were measured, allowing assessment of associations with disease status as well as autoantibody burden. To our knowledge, this is among the first studies from India to jointly examine DII, DDS, and MPA in relation to RA and RA-related autoantibodies in a case-control design.

Despite its strengths, this study has several limitations. First, although the study was designed as a matched case-control study (with age/sex frequency matching at recruitment), significant differences in age and BMI remained between cases and controls, and unconditional logistic regression was used because complete matching was not achieved. This may have introduced residual imbalance and affected comparability between groups. In addition, because recruitment of healthy relatives was not feasible, controls were selected from schools, institutions, and private organizations in Hyderabad, which may have contributed to the younger age profile of controls. Second, dietary intake was assessed retrospectively using an FFQ, including recall of pre-disease dietary habits. This introduces the possibility of recall bias, response error, and exposure misclassification (including DII), particularly if cases recalled past diet differently after diagnosis. Although the FFQ was validated for the local population and interviewer-administered, some misclassification cannot be excluded. Third, the case-control design precludes causal inference and does not establish temporality; although we recruited newly diagnosed RA cases to reduce this concern, reverse causation remains possible (e.g., symptom-related changes in appetite or food choices before diagnosis). Fourth, residual confounding is a major limitation because data were not available for several important factors associated with both diet and RA risk, including socioeconomic status/wealth, education, physical activity, smoking, alcohol use, medication use (e.g., NSAIDs/corticosteroids before diagnosis), and genetic susceptibility. Fifth, the study was conducted in an urban population in southern India, and the predominance of one sex in the sample may limit generalizability to other settings and populations. Sixth, The DII was calculated using 36 of the 45 original food parameters. Because several parameters, particularly anti-inflammatory components, were unavailable, the calculated DII scores may have been shifted toward a more pro-inflammatory distribution, limiting direct comparison of absolute DII values with studies using a more complete set of DII components [33,34,36].

Similarly, MPA was calculated using only 10 micronutrients because the required EAR/CV inputs were available only for these nutrients. For several micronutrients, median PA values were close to zero in both groups, likely reflecting a floor effect of the EAR-based probability approach rather than absence of between-person variation in FFQ-estimated intake; therefore, micronutrient-specific PA medians were interpreted cautiously, with greater emphasis on comparative analyses, MPA, and complementary indicators (DDS and DII). The reported PAF estimates should be interpreted as theoretical because, in an observational case-control study, they depend on strong causal assumptions and should not be considered direct estimates of preventable disease burden.

## CONCLUSION

In this case-control study, RA cases had poorer dietary quality, lower dietary diversity, lower micronutrient adequacy, and a more pro-inflammatory dietary profile compared with healthy controls. Lower DDS and MPA, and higher DII scores, were associated with higher odds of RA. Prospective and intervention studies are needed to confirm temporality and assess whether improving dietary quality and reducing dietary inflammatory potential can contribute to RA prevention and management.

## Data Availability

All data produced in the present study are available upon reasonable request to the Corresponding author.

## Acknowledgment

The authors gratefully acknowledge Dr. Bhagavathi, VNL Project Consultant (Ayurveda), for support in subject recruitment, and Ms. Neha Lodaya, M.Sc. (Intern), SVT Mumbai, for assistance with diet data curation.

## Data Availability Statement

The raw data is available with Dr. Devraj Parasannanavar and would be made available to anyone upon written request with justification.

---

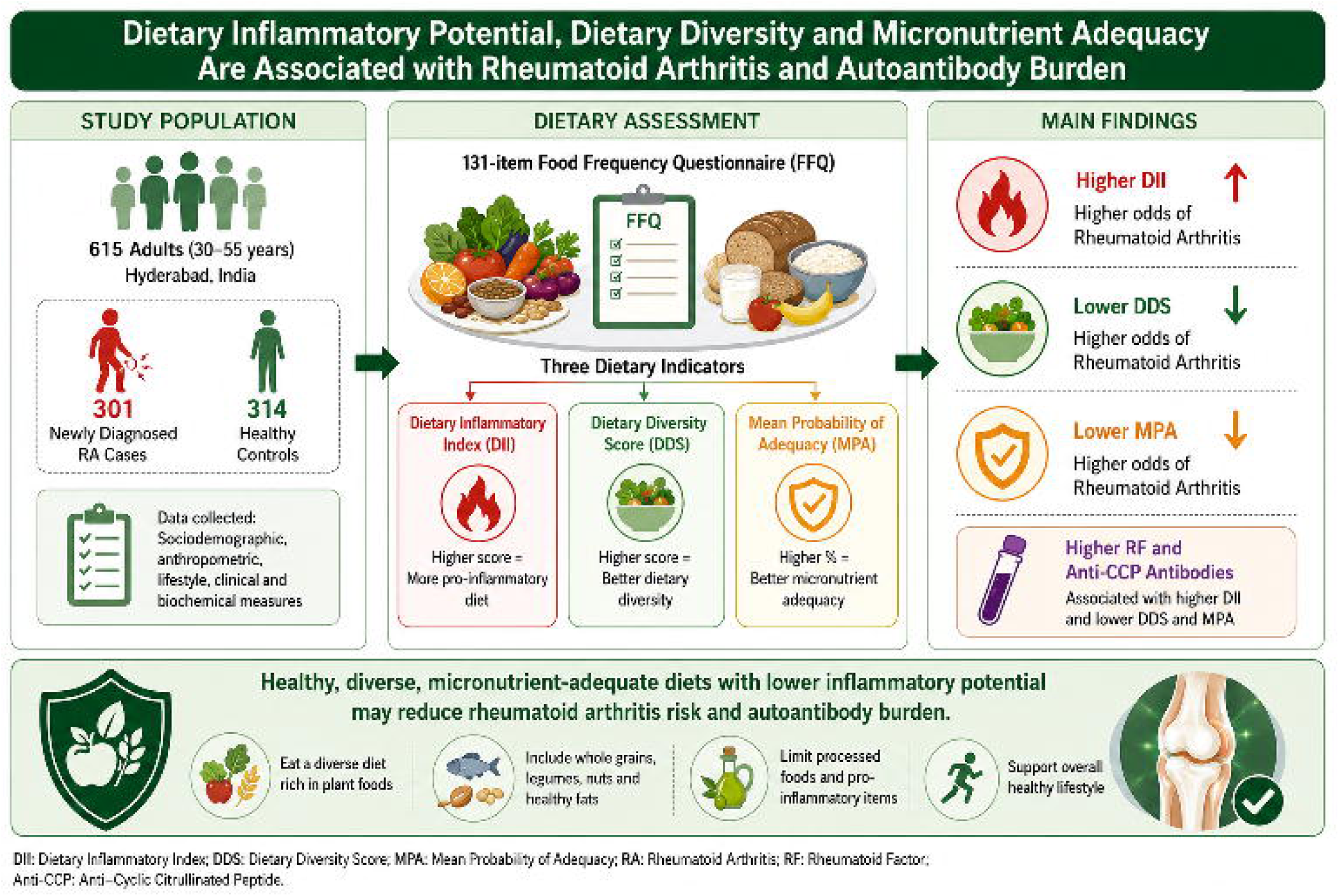

