## Supplementary Appendix for "Dietary Inflammatory Potential, Dietary Diversity and Micronutrient Adequacy Are Associated with Rheumatoid Arthritis and Autoantibody Burden: A Case-Control Study from India"

**Supplementary Table 1**. Included and Excluded Parameters to calculate Dietary Inflammatory Index

| | |
|---|---|
| Included Parameters | |
| Energy | Iron |
| Protein | **Zinc** |
| Carbohydrates | **Thiamine** |
| Fats | **Riboflavin** |
| **MUFA** | Vitamin B12 |
| **PUFA** | **Vitamin C** |
| Trans fats | **Vitamin A** |
| **Fiber** | **Vitamin D3** |
| **Folic acid** | **Vitamin B6** |
| **β-carotene** | Total cholesterol |
| Vit E | **Magnesium** |
| Niacin | **Selenium** |
| **Garlic** | **n-3 Fatty acids** |
| **Ginger** | **n-6 Fatty acids** |
| **Turmeric** | **Onion** |
| **Flavanones** | Saturated Fat |
| **Isoflavones** | **Pepper** |
| **Flavones** | **Flavan-3-ol** |
| Excluded Parameters | |
| **Alcohol** | **Saffron** |
| **Caffeine** | **Green/black tea** |
| **Eugenol** | **Anthocyanidines** |
| **Flavonols** | **Rosemary** |
| **Thyme/oregano** | |

Food parameters highlighted in bold are assumed to have an anti-inflammatory effect score

**Supplementary Table 2**. Population Attributable Fractions (PAFs), Exposure Prevalence, and Adjusted Odds Ratios for Incident Rheumatoid Arthritis

| Exposure (risk category vs reference) | Reference category | Prevalence of exposure (Pe) | Adjusted OR (95% CI) | PAF (%) | 95% CI |
|---|---|---|---|---|---|
| DII (T2+T3 vs T1) | T1 (lowest tertile) | 66.70% | 2.17 (1.49–3.16) | 43.8 | 24.6–58.0 |
| DDS (Low <5.0 vs High ≥5.0) | High DDS (≥5.0) | 37.60% | 2.70 (1.90–3.90) | 39.5 | 25.3–49.9 |
| MPA (Inadequate <50% vs Adequate ≥50%) | Adequate intake (≥50%) | 71.90% | 1.84 (1.24–2.72) | 37.7 | 14.7–54.6 |

OR = Odds Ratio adjusted for covariates (specify your model variables if needed); Pe = prevalence of exposure in the study population; PAF = Population Attributable Fraction calculated assuming a causal relationship; DII = Dietary Inflammatory Index; DDS = Dietary Diversity Score; MPA = Mean Probability of Adequacy; T= Tertile;
*Unconditional Logistic Regression models* were adjusted for age, sex and body mass index;
